# Auditory deprivation during development alters efferent neural feedback and perception

**DOI:** 10.1101/2022.11.24.22282369

**Authors:** Srikanta K Mishra, David R. Moore

## Abstract

Auditory experience plays a critical role in hearing development. Developmental auditory deprivation due to otitis media, a common childhood disease, produces long-standing changes in the central auditory nervous system, even after the middle ear pathology is resolved. The effects of sound deprivation due to otitis media have been mostly studied in the ascending neural system but remain to be examined in the descending pathway that runs from the auditory cortex to the cochlea via the brainstem. Alterations in the efferent neural system could be important because the descending olivocochlear pathway influences the neural representation of transient sounds in noise in the afferent auditory system and is thought to be involved in auditory relearning following injury. The main objectives of the present study were to (1) investigate whether degraded auditory input due to otitis media during childhood is associated with weakened medial olivocochlear efferent neural responses, even after the resolution of the middle ear pathology, and (2) to examine the involvement of the efferent neural feedback in perceptual masking deficits associated with auditory deprivation due to otitis media.

We measured contralateral inhibition of otoacoustic emissions—a biomarker for medial efferent activity—and speech-in-noise recognition in children with a medical history of otitis media (N=76) and age-matched controls (N=99). All children had normal auditory function at the time of experimentation.

We found that the inhibitory strength of the medial olivocochlear efferents is weaker in children with a documented history of otitis media relative to controls. In addition, children with otitis media history required a more advantageous signal-to-noise ratio than controls to achieve the same criterion performance level. Importantly, the deficits in perceptual masking were related to efferent inhibition, and these effects could not be attributed to the middle ear or cochlear mechanics.

These findings raise the possibility that perceptual masking deficits—a hallmark of impaired (central) auditory processing—resulting from otitis media can arise from the altered brainstem efferent feedback. To date, it was known that degraded auditory experience reorganizes the ascending neural pathways; here, we show that the lack of optimal auditory input to the afferent system during development could have a long-standing impact on the functioning of the descending neural pathways.

## Introduction

Both good and bad auditory experiences can alter hearing. Although varying degrees of malleability in auditory structure and function are observed across the lifespan, the auditory system has remarkable plasticity during early development (e.g., for reviews^1,2^). For example, Buran *et al*.^3^ showed that inducing conductive hearing loss (CHL) produced a greater deficit in frequency modulation detection in juveniles than in adult animals.

Conductive hearing loss due to otitis media (OM) is a useful model for understanding how reversible developmental auditory deprivation affects sound processing in the central auditory nervous system. For example, bilateral CHL has been shown to disrupt the temporal response properties of auditory cortex neurons^4,5^. OM is the most common reason for children to go to the doctor. Although recent epidemiologic data suggest a decline in the incidence of OM since the mid-90s, the condition is still remarkably prevalent, affecting at least 70% of children in the first 5 years^6,7^. OM can attenuate the overall amplitude of the acoustic signal and alter the temporal fidelity of the input signal to the cochlea by delaying the transmission of low-frequency sounds^8–10^. Several animal and human studies show diminished and temporally degraded sound-evoked activity produced by OM, affecting neural representations of acoustic stimuli and perceptual ability during and after the resolution of the middle ear pathology (for review^11^). A general hypothesis is that diminished perceptual skills result from altered sound encoding in the central auditory system, for example, through loss of synaptic inhibition^12^. However, how the degraded auditory experience due to OM causes enduring perceptual deficits or the specific neural mechanisms underlying the deficits are not fully understood.

Most human studies show that temporary CHL due to OM is associated with a reduced ability to understand speech in the presence of background noise, even without elevated pure tone sensitivity. This perceptual effect of OM has been observed across multiple speech stimuli (words or sentences), masker types (speech-shaped noise or competing talkers), and methods of measurement [e.g., adaptive vs. fixed signal-to-noise ratio (SNR)]^13–17^. However, the neural substrate of deficits in perceptual masking is not yet clear. Some psychophysical evidence suggests impaired temporal resolution as a contributing factor for abnormal speech-in-noise recognition in children with an OM history^14,18–22^. Studies using click-evoked auditory brainstem responses show delayed wave III and/or V latencies and increased interwave intervals, indicative of immaturity in neural conduction in children with an OM history despite the resolution of the middle ear fluid^23–26^. However, only one study linked this abnormal brainstem physiology with binaural hearing deficits as measured by masking level difference^23^, while another study did not find a significant association between delayed brainstem responses and speech-in-noise measures^25^.

An important mechanism that enhances the neural representation of transient sounds in the presence of background noise is the efferent auditory pathway. Specifically, the medial efferents descend from the superior olivary complex and project onto the cochlear outer hair cells, forming the last leg of the descending neural pathway, which originates from the auditory cortex (for review^27^). The medial olivocochlear efferents reduce the gain of the cochlear amplifier and inhibit the auditory nerve response to background noise; this action partially restores the dynamic range of the neural response to rapidly changing sounds or acoustic transients^28–30^ (for review^31^). In humans, otoacoustic emissions (OAEs) can be applied to measure the functioning of the brainstem efferent feedback circuitry, also called efferent inhibition, efferent neural inhibition, or efferent strength^31^. The efferent unmasking mechanism is robust in the juvenile population, enabling tone discrimination in noise and word-in-noise recognition^32,33^. In children, medial olivocochlear efferent fibers can also modulate peripheral frequency selectivity, that is primarily determined by outer hair cells^34^.

Evidence from human studies suggests that the strength of the efferent unmasking mechanism can be influenced by auditory training and musical experience^35–37^ (for review^38^). However, the relationship between efferent dysfunction and degraded auditory experience during development, and its impact on perceptual deficits, remains to be elucidated. In a mature mouse model, Liberman *et al*.^39^ reported reduced lateral (but not medial) olivocochlear efferent innervation and a corresponding loss of afferent synapses in ears with CHL, compared to control ears, ∼ 1 year from a tympanic membrane resection. Although the extent to which this neural structural loss would affect auditory neurophysiology or function is not known, their study supports the growing evidence for the importance of descending neural control of the cochlea in shaping the afferent neural pathway.

In this study, we asked in children whether the perceptual deficits associated with auditory deprivation due to OM are related to the strength of the medial efferent feedback pathways to the cochlea. We hypothesized that degraded auditory signal transmission through the developing auditory system alters the strength and functioning of the efferent system. Weakened efferent neural feedback would affect the ability to discriminate sounds in noise. To address the research question, we measured speech-in-noise recognition and sound-evoked, medial efferent inhibition of OAEs in controls and children with documented medical histories of degraded auditory experience due to OM. Both groups had normal peripheral auditory function at the time of testing, as indexed by hearing thresholds, wideband absorbance, and OAEs.

## Materials and methods

### Participants

A total 151 children were enrolled in this study (mean age= 8.1 years; range= 5 to 12.8 years; girls= 76). All children had 20 dB HL or better hearing thresholds at octave frequencies from 250 through 8000 Hz and normal, type-A tympanograms^40^. In addition, the wideband absorbance, measured using a HearID system (Mimosa Acoustics, Champaign, IL), for every child was within the reported normative range^41^. No children had clinical histories of childhood communication or behavioral disorders for which they were receiving an intervention.

A Control group (n= 99, girls= 48) had no significant history of middle ear disease, i.e., two or fewer documented OM episodes since birth and no documented OM in the previous one year. An OM group (n= 52, girls= 28) had documented medical history of bilateral OM with flat or type B tympanogram and CHL. The last episode of OM occurred at a (mean) age of 3 years (range= 2 to 4 years). The worst hearing thresholds ranged from 25 to 45 dB HL (mean= 31.3) at the time of the last OM episode (Figure 1A). The middle ear and hearing history for both groups were collected from clinical records. The information regarding the last OM episode is an approximation and has the inherent assumption that the OM episode after the last episode, if any, was not significant enough for a clinic visit. All test procedures were conducted in a sound-booth. The ear with a lower hearing threshold at 1000 Hz or the right ear, if thresholds were the same, was selected for running experimental procedures.

**Figure 1.**
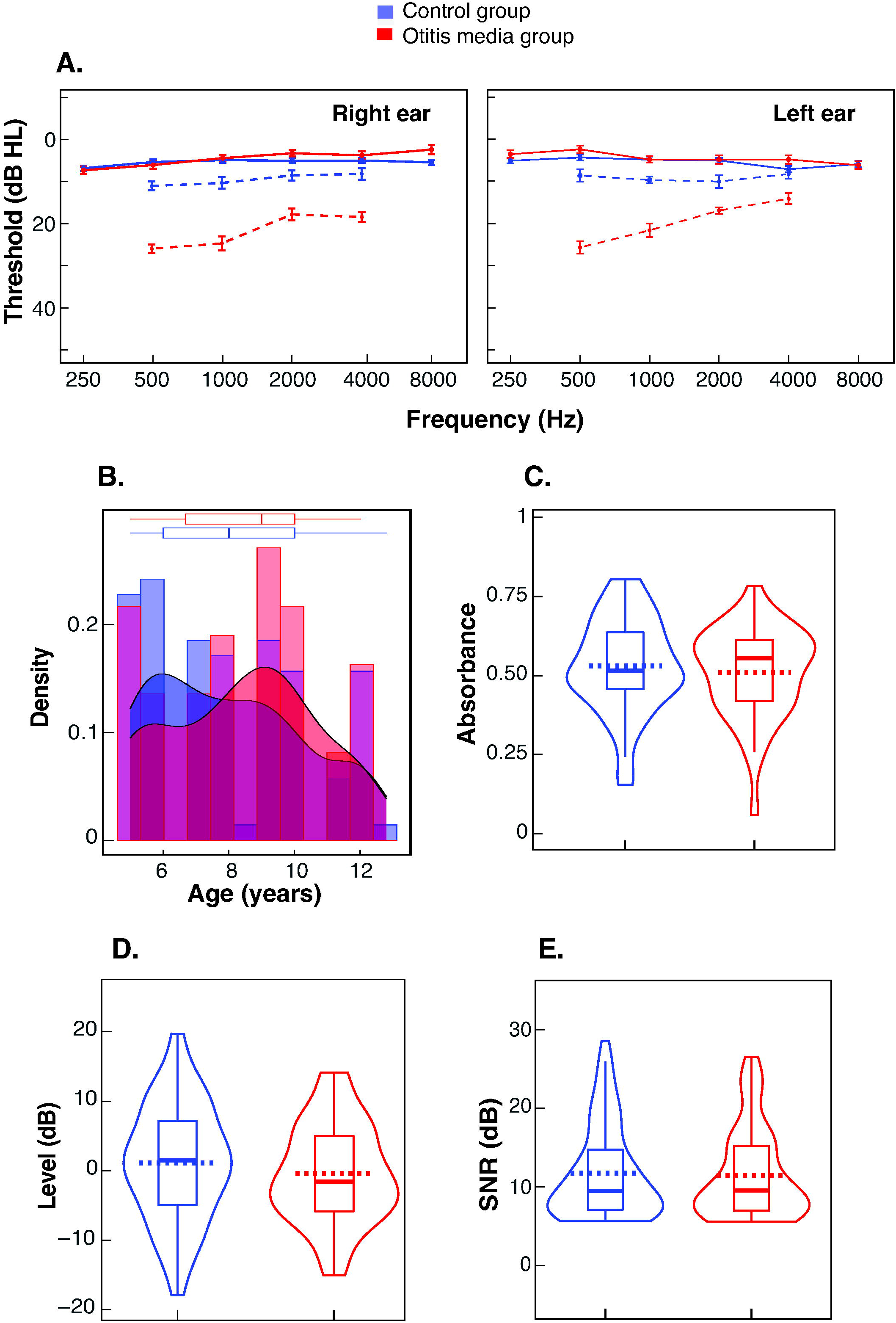
Auditory function in the control and otitis media groups: **A**. Mean thresholds for the control and OM groups plotted separately for right and left ears; error bars show SD. Solid lines represent thresholds at the time of testing from all children, and broken lines show thresholds at the last OM episode (n= 16; mean age= 3 yrs) and an equivalent time for controls (n= 14). **B**. Density plot for age distribution for the two groups with box-whisker plots on the top. **C**. Violin plots for absorbance for the control and OM groups. **D**. Violin plots for SFOAE level, and **E**. SFOAE signal-to-noise ratio for the control and OM groups. For all violin plots, violins show kernel probability density, boxes are interquartile ranges (with median and mean as solid and broken lines, respectively), and whiskers are 1.5 times the interquartile range.

### Stimulus frequency otoacoustic emissions (SFOAEs) and contralateral stimulation

SFOAEs, recorded at low probe levels, optimally represent cochlear amplifier gain^42^. In addition, among all OAE types, SFOAEs are more appropriate for measuring efferent inhibition^43^. Considering the non-frequency specific nature of medial efferent innervation of the cochlea^44^, the efferents will be acting along the length of the basilar membrane when inhibition of SFOAEs is observed for the 1000 Hz band. Therefore, contralateral inhibition of SFOAEs at 1000 Hz center frequency can be considered as a marker of the efferent neural response across the entire cochlea^45^.

The equipment set-up and calibration for recording swept-tone SFOAEs are previously described by Mishra and Talmadge^46^, The protocol for contralateral stimulation was similar to a previous study^47^. Stimulus generation and recording of signals from the ear canal were performed using the ER-10B+ microphone probe assembly (Etymotic Research, Elk Grove Village, IL). They were controlled via a MOTU 828x audio interface (MOTU, Cambridge, MA) using custom-built programs on the RecordAppX software ^48^.

The target frequency range for recording SFOAEs was 707 to 1414 Hz to estimate efferent inhibition at 1000 Hz center frequency. This center frequency has a high SNR for SFOAEs^46,49^, shows robust efferent effects^50^ and is free from short-latency components for the probe level^51^. SFOAEs were measured via a suppressor paradigm using 40 dB probe and 60 dB suppressor tones. The suppressor frequency was 1.1 times higher than the probe, and both tones were swept from low to high at 0.188 octave/s. The phase was inverted for every other use of the suppressor.

SFOAEs were recorded with and without continuous broadband noise presented at 60 dB SPL to the contralateral ear using an ER-2 insert phone. The contralateral interval had a 2s lead and was interleaved with the probe sweep, with a 3s gap between two successive sweeps. The child sat on a recliner and was instructed to remain calm and quiet. Children were also offered an age-appropriate, close-captioned video if they wanted to watch it.

Estimates of magnitude, phase, and noise floor were obtained using a least-squared filter modeling^46^. SFOAEs were estimated by minimizing the sum of the squared error between the model and the response. The noise floor was the average pairwise sweep difference between the probe-plus-suppressor and the probe alone. The efferent inhibition (%) was computed using a vector difference method that considers the SFOAE phase and normalizes the magnitude to the baseline SFOAE to compute the efferent inhibition for OAEs with SNR≥ 6 dB^52^. The absence of middle ear muscle reflex effects was ensured by using a group delay test with a 4ms criterion^52^. The SNR criterion was not met in four children, and data from two children failed the group delay test as well as the SNR criterion.

### Hearing in Noise Test for Children (HINT-C)

Speech recognition threshold (SRT) was measured monoaurally using the American HINT-C^53^. The HINT-C is based on 13, ten-sentence lists that are phonemically balanced. The test was implemented in a Windows laptop with a USB sound card (Scarlett 2i2, Focusrite, UK) and TDH39P headphones. The speech and noise were co-located in the front. The level of noise, which is matched to the long-term-average spectrum of the sentences, was fixed at 65 dB. The speech level was varied adaptively to achieve the target SNR. The starting SNR was 10 dB with an initial step size of 5 dB for the first four trials, reducing to 3 dB. Each run included two distinct sentence lists. The child was instructed to repeat the entire sentence correctly. Scoring was based on correctly reported complete sentences allowing minor variations in articles and verb tenses. The SRT is defined as the mean of SNRs from the fifth through the 21st trials. The 21st sentence is not presented, but its SNR is predicted from the previous response.

Each child was presented with a practice run for familiarization, followed by two test runs. Thresholds from those two runs were averaged to get an estimate of SRT. If the SRTs between two runs differed by more than 10%, a third run was used, and the two lowest scores were averaged. The participants were native speakers of American English. Only 100 (controls= 65) out of 151 children completed the HINT-C. Notably, 19 (controls= 14) out of 46 (controls= 33), 5-to-6-yr-olds could reliably complete the test.

### Statistics

Statistical analyses were conducted using Jamovi software (v 2.3.18). Group differences (control and OM) were compared using Welch’s t-tests. Analysis of covariance (ANCOVA) with age (log-transformed) as a covariate was conducted to compare group differences for efferent inhibition and SRT. Effect sizes (*η* ^2^) for ANCOVA were interpreted according to Mile and Shevlin^54^. Linear regression was used for modeling efferent inhibition and SRT. All reported model predictors have variance inflation factors lower than 3. An effect was considered significant if *p*≤ 0.05. Missing data were not imputed.

### Data availability

The data and custom analysis scripts that support the findings of this study can be made available upon request to the corresponding author.

## Results

### Peripheral auditory characteristics at the time of testing

Hearing sensitivity, middle ear function, and cochlear function were normal in all children at the time of testing (Figure 1). There was no significant age difference between the control and OM groups (control: mean= 8.02 yrs, SD= 2.30; OM: mean= 8.40 yrs, SD= 2.23; Welch’s *t*_106_ = −0.99, *p*= 0.32). Hearing thresholds for all audiometric frequencies were ≤ 20 dB HL for all children, and there were no significant group differences for any frequencies or ears (F_1,149_ = 1.33, *p*= 0.25). Wideband absorbance did not differ between the groups (control: mean= 0.53, SD= 0.15; OM: mean= 0.51, SD= 0.14; Welch’s *t*_77_ = 0.69, *p*= 0.49). For brevity and relevance to efferent measurements, absorbance results are presented for 1000 Hz center frequency with one-octave band; however, the two groups did not differ significantly across the measured frequency range (250-6000 Hz). The mean and range of variation in wideband absorbance across children in both groups are consistent with a previous normative study^41^.

SFOAE data from six children in the control group did not meet the SNR criteria. For the remaining children, the mean SFOAEs were not significantly different across groups (Figure 1D; control: mean= 1.12, SD= 8.28; OM: mean= −0.41, SD= 7.09; Welch’s *t*_120_ = 1.17, *p*= 0.24). Likewise, there was no significant difference in SNR for SFOAEs between the two groups (control: mean= 11.78, SD= 9.50; OM: mean= 11.49, SD= 9.56; Welch’s *t*_108_ = 0.28, *p*= 0.78).

### Effect of OM history on efferent inhibition and SRT

Efferent inhibition (Figure 2A) for the OM group was significantly lower than for the Controls (ANCOVA with a fixed factor of group and age as a covariate; F_1,142_ = 7.11, *p*= 0.01, *η*^2^ = 0.05). However, the effect size was small. No significant effect of age was observed (age: F_1,140_ = 0.61, *p*= 0.44). Although no significant difference between the two groups in middle ear transmission (absorbance) or cochlear function (SFOAE) was observed, linear regression with wideband absorbance, SFOAE level, and SNR was conducted to ensure that these factors did not contribute significantly to the efferent inhibition. The model was not significant (F_3,96_ = 0.36, *p*= 0.77, R^2^ _adjusted_ = −0.02), showing that neither the middle ear nor outer hair cells functioning at the time of testing could explain the observed efferent inhibition effects of OM.

**Figure 2.**
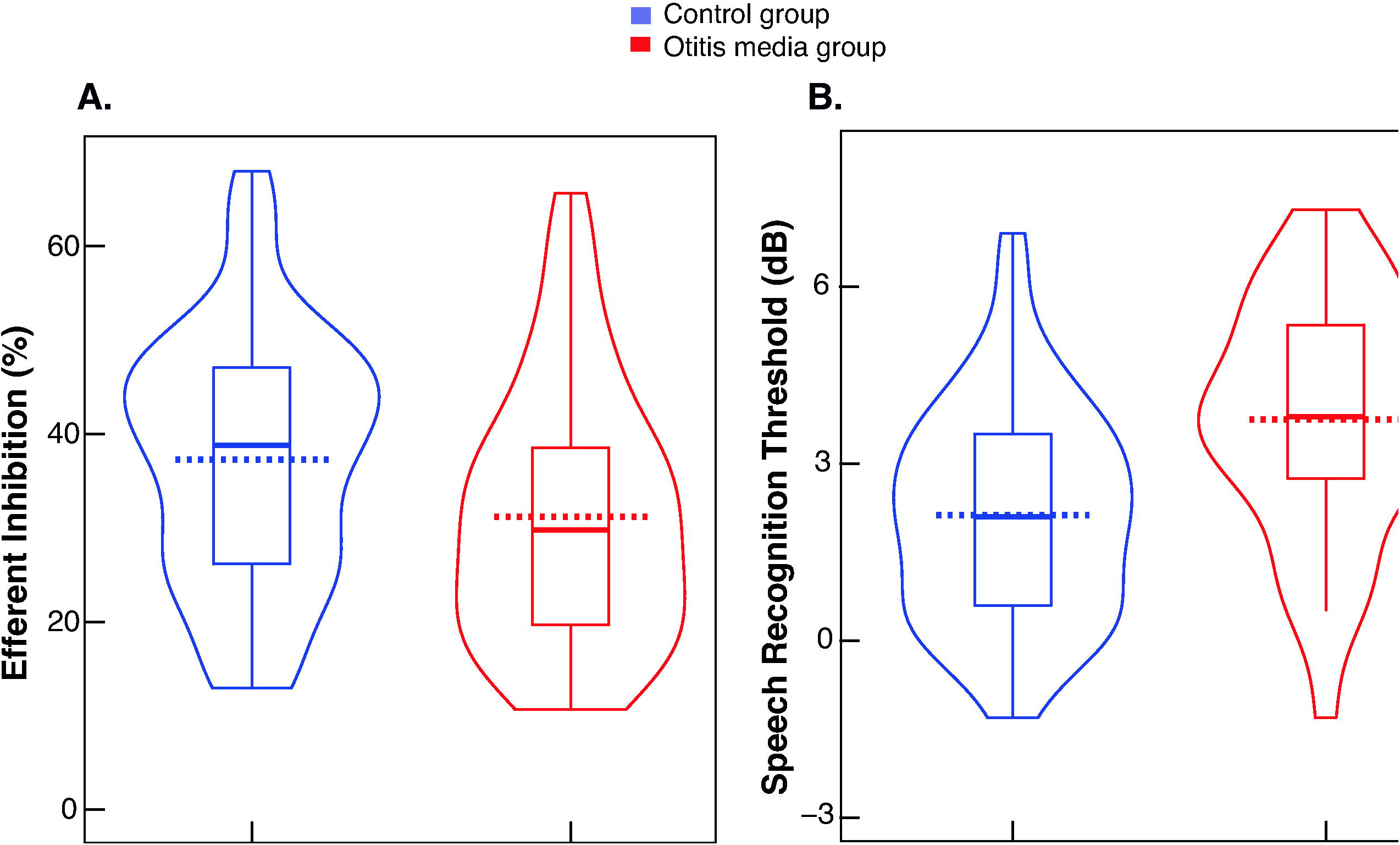
Efferent neural inhibition and perceptual measures: **A**. Violin plot of efferent inhibition for control and OM groups. **B**. Violin plot of speech recognition threshold for control and OM groups. Violin plots show kernel probability density, boxes are interquartile ranges (with median and mean as solid and broken lines, respectively), and whiskers are 1.5 times the interquartile range.

The OM group had higher (poorer) SRT relative to controls (Figure 2B), after controlling for age differences (ANCOVA: Group: F_1,97_ = 30.2, *p*< 0.001; *η*^2^ = 0.17; age: F_1,97_ = 50.5, *p*< 0.001; *η*^2^ = 0.28), with medium effect sizes. The improvement in SRT with age did not differ statistically between groups, as the interaction between age and group was not significant (Figure 3A; ANCOVA: Age×Group: F_1,96_ = 0.0001, *p*= 0.99).

**Figure 3.**
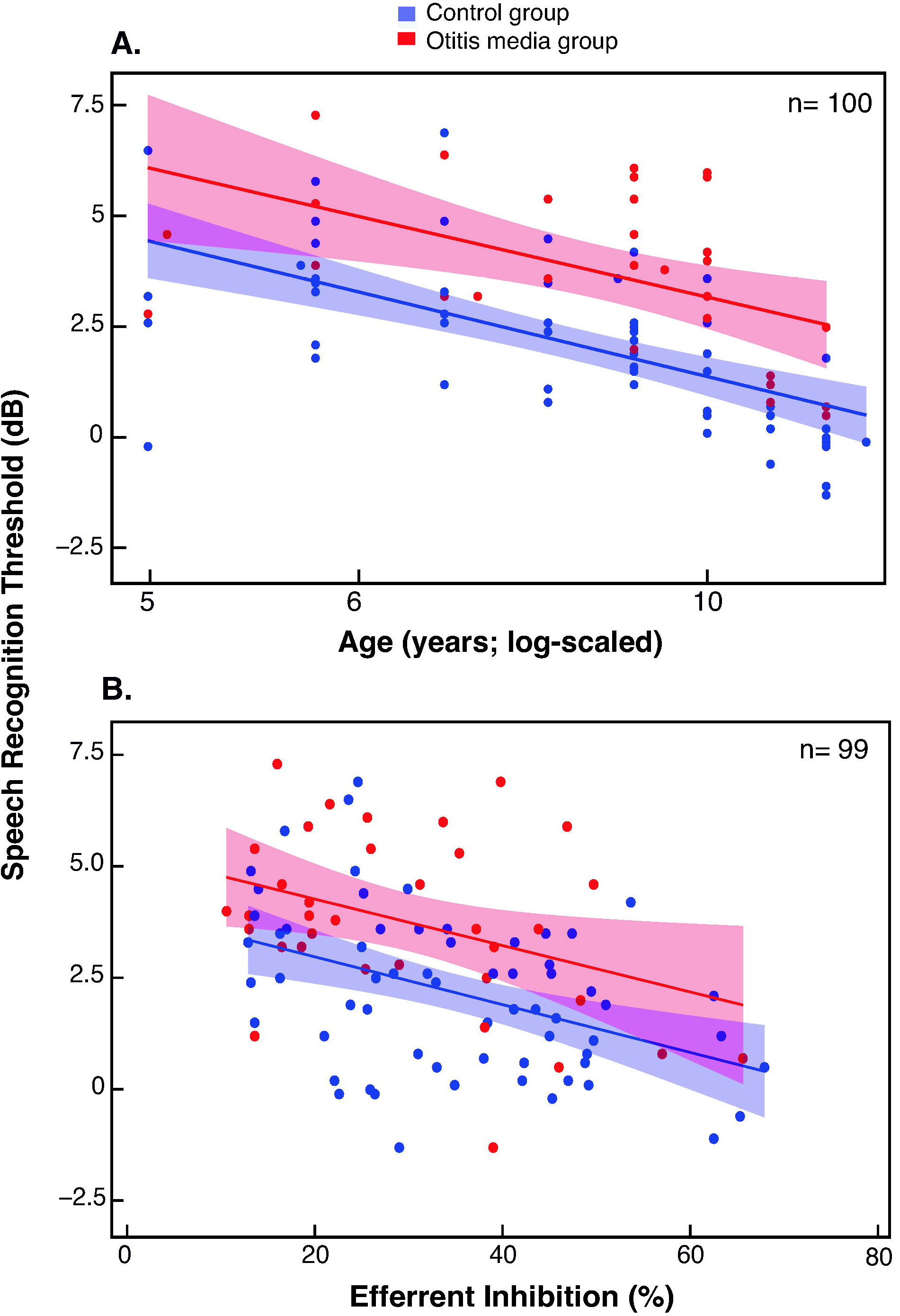
The relationship between efferent inhibition and perceptual masking: **A**. Speech recognition threshold as a function of age with 95% confidence intervals as shaded areas, blue and red for the control and OM groups; **B**. Speech recognition threshold as a function of efferent inhibition with 95% confidence intervals as shaded areas, blue and red for the control and OM groups based on the linear regression model outcomes (Table 1).

To examine whether the duration of normal auditory experience following resolved OM is related to efferent inhibition and SRT, separate partial correlations were conducted, controlling for age. The normal duration following resolved OM (mean= 5 yrs, range= 1 to 9) was significantly related to the SRT (data not shown) but not the efferent inhibition (Figure 3B: *r*= 0.07, *p*= 0.31, n= 51; SRT: *r*= −0.35, *p*= 0.02, n= 35).

### Modeling speech-in-noise recognition

Linear regression with predictors age, absorbance, SFOAEs, efferent inhibition, group (OM and control), and sex was conducted to model the combined effects of factors on speech-in-noise recognition. The model was significant (F_6,66_ = 12.9, p= <0.001, R^2^ _adjusted_ = 0.49), and the model outcomes are presented in Table 1. The regression model explained 49% of the variance observed in the SRT. Age, efferent inhibition, and group significantly contributed to the model. The SRT improved (decreased) with age and efferent inhibition, and declined (increased) with OM history. Middle ear, cochlear functioning, and sex effects were not significant. Figure 3B plots the relationship between efferent inhibition and SRT.

## Discussion

Several studies showed poorer signal detection and discrimination in noise following OM even after the middle ear disease is resolved^13–17,55–57^. The neural mechanism by which auditory deprivation associated with OM increases the vulnerability to perceptual masking is not fully understood, although it is generally attributed to the neural reorganization in the ascending auditory pathway (for review^11^). In the present study, we found significantly lower efferent inhibition and poorer speech-in-noise recognition in children with documented OM history relative to controls, even when the peripheral auditory function was normal and indistinguishable from controls. In addition, poorer speech-in-noise recognition in children with OM history was related to efferent neural feedback. These findings suggest that compromised afferent input due to OM during development produces long-term changes in the medial efferent feedback, which has perceptual consequences. The current results provide new evidence that medial olivocochlear efferent dysfunction associated with OM history can lead to behavioral hearing deficits, specifically, increased vulnerability to perceptual masking.

### Methodological considerations

A potential limitation of using clinical records is that the number of OM episodes may not be precise, which requires a longitudinal design. For example, Halliday and Moore^58^ found that around half of the studied sample (*n*= 112) had frequent episodes of OM in one or both ears; however, only a small portion of this would have been detected through a visit to health care providers. As a result, the OM group in this study may represent children with more frequent OM episodes who received medical intervention. In fact, the threshold data at the last OM (Figure 1A) are consistent with the audiometric profile of the high-prevalence OM group reported by Hogan and Moore^59^ (their Table 1). A related issue is that threshold test data was not available for all episodes of OM, although thresholds from the last OM episode were available. As such, even in a longitudinal design, it may not be feasible to measure behavioral thresholds reliably in infants and toddlers with OM, and when the measurements are feasible, the test results may not be very sensitive to fluctuations that can occur with OM. Further, only about 60% of children with OM experience mild-to-moderate CHL^60^. Whitton and Polley^11^ claimed that CHL, not the mere presence of OM, is the determining factor for perceptual and physiological alterations in the auditory system. However, findings from a recent study refuted this claim for a tone-in-noise detection task^61^.

### Experience-dependent changes in the efferent neural pathway

The corticofugal fibers originating from neurons in the auditory cortex exert control on the periphery via the olivocochlear efferents (for review^62^). Specifically, the olivocochlear efferent neurons modulate the afferent neural representation of the acoustic input. Evidence from mature animal models suggests that the brainstem efferent feedback circuitry is critically important for relearning sound localization following injury^63^. However, very few studies have investigated the plasticity of the efferent neural system. Kraus and Illing^64^ showed that medial (not lateral) olivocochlear neurons are the major source of synaptic reorganization in the ventral cochlear nucleus after cochleotomy in adult rats. In human adults, de Boer and Thornton^35^ have shown that efferent neural strength and associated perceptual masking can be improved through targeted auditory training. Until recently, it was not known whether sound deprivation due to CHL alters olivocochlear efferents. Liberman *et al*.^39^ showed degeneration of the lateral efferent terminals following chronic conductive hearing loss in adult mouse models. However, the functional consequences of alterations in lateral olivocochlear neurons are less known. We have carefully eliminated middle ear and cochlear factors in the efferent inhibition measurements, and demonstrated that children with OM in the first three years of life have weaker medial efferent inhibition relative to controls. Similar findings in humans have been reported for the acoustic reflex—another brainstem feedback pathway^25^.

The lack of an age effect on efferent inhibition for children is not surprising, given that the medial efferent pathway is considered to be mature at full-term birth^65,66^. In addition, we found no relationship between normal auditory experience and efferent inhibition in the OM group, suggesting that efferent dysfunction associated with temporary CHL in early childhood (< 3 yrs) may not recover to normalcy even after 5 years of typical auditory experience.

The efferent control of the cochlear mechanics is reasonably well understood to predict the potential consequences of CHL. Theoretically, CHL, due to OM, would attenuate the sound level reaching the cochlea. As a result, the medial olivocochlear neurons will not receive optimal stimulation at a sound level for which the efferent action is known to be effective^43,67^. This may temporarily shift the dynamic range of the efferent neural response. Such altered efferent biomechanics during the early developmental period may take a longer recovery time or may not restore naturally after the OM is resolved. However, the reduced efferent inhibition could potentially be restored via targeted auditory training^35^.

### Medial olivocochlear feedback as a mechanism for perceptual consequences of OM

Despite firm evidence of perceptual masking deficits due to auditory deprivation associated with OM history, the underlying neural mechanisms are less clear. Few previous studies applied both neural/physiologic as well as perceptual measures for studying the neural substrate for poorer perceptual skills in children with OM history. Hall and Grose^23^ reported significant correlations between interaural asymmetries of the interwave intervals of auditory brainstem responses and the masking level difference. Their findings suggested a link between abnormal brainstem processing and binaural hearing deficits in children with OM history. We selected speech stimulus instead of tones as the effects of OM history for speech have been consistently demonstrated. Simpler tones may not reveal the perceptual deficits associated with auditory deprivation due to OM, even when OM causes CHL^61^. Children with OM history required a more favorable SNR (∼ 2 dB) than controls to achieve the same criterion performance level. We have demonstrated that medial efferent inhibition, a brainstem neural substrate that supports the unmasking of transient sounds in noise, predicts speech-in-noise recognition for the OM group. This finding supports reduced efferent feedback as a mechanism for poorer signal discrimination in noise for children with OM history. However, chronological age, efferent inhibition, and OM history combined could explain only 50% of the variance observed in the SRT. This suggests that additional factors, such as central (afferent) auditory and cognitive processes, are involved in poorer perceptual masking in children with OM history.

The time course of the recovery of the neurophysiologic and perceptual effects of OM is unknown. However, the significant relationship between the time since resolved OM and SRT suggests that typical auditory experience (∼ 5 yrs) following OM may slowly restore perceptual masking. The improvement in SRT but not efferent inhibition with the normal auditory experience period following resolved OM may suggest that children learn coping strategies and/or rely on additional mechanisms to filter out noise from the speech.

### Translational significance

Findings from the present study have implications for understanding the role of the efferent neural system in human hearing and related clinical applications. First, efferent inhibition may be reduced in other forms of developmental deprivation, such as minimal hearing loss and extended high-frequency hearing loss, which are associated with OM history^68^. It could contribute to the perceptual deficits observed in such sub-clinical disorders. Second, medial olivocochlear efferents may play a role in preventing noise-induced hearing damage^69^. Since children with OM history have a weakened efferent system, they may be particularly vulnerable to future noise-induced cochlear damage or synpatopathy. Third, children with stronger medial efferent feedback may have relatively lesser consequences from auditory deprivation due to OM. This prediction is consistent with the role of efferents in hearing (perceptual and physiologic) development and plasticity^32,63,70^. Related to this, enhancing the medial efferent feedback via pharmacological treatment or targeted auditory training could be a viable strategy to rescue and restore afferent neural encoding of sounds and minimize the long-term perceptual consequences of OM. Finally, medial efferent inhibition shows large inter-individual variability in human adults and children^32,71,72^. Typically, the OM history is not considered in studies investigating medial efferent reflex in humans; instead, peripheral auditory functioning is measured. Considering the deficiencies in the efferent functioning associated with OM history despite normal peripheral auditory function, we speculate that OM history may partly explain the variability in the efferent inhibition in normal-hearing individuals.

The mean SRT was ∼ 2 dB poorer for children with OM history than controls, which corresponds to a ∼15-percent deficit in speech-in-noise recognition scores according to psychometric functions measured in children, albeit for digits^73^. This perceptual deficit is highly relevant for classroom environments where background noise levels are often high^64^ and may impact learning in the classroom.

## Conclusions

Children with OM history had significantly poorer SRTs and reduced efferent inhibition relative to Controls. In addition, medial efferent inhibition predicted the ability to understand speech in the presence of noise. These effects suggest that the lack of optimal auditory experience due to OM produces long-standing effects on efferent neural feedback. Importantly, it also suggests medial efferent inhibition as a neural correlate of the perceptual masking deficits observed in children with OM history. The current study demonstrates dysfunction in the brainstem neural feedback circuitry as a long-term consequence of OM complementary to an otherwise extensive literature that predicts perceptual effects of auditory deprivation due to central auditory degeneration.

## Data Availability

All data produced in the present study are available upon reasonable request to the authors

## Abbreviations

ANCOVA: Analysis of Covariance
CHL: Conductive Hearing Loss
dB: Decibel
HINT-C: Hearing in Noise Test for Children
HL: Hearing Level
OM: Otitis Media
SD: Standard Deviation
SFOAEs: Stimulus Frequency Otoacoustic Emissions
SNR: Signal to Noise Ratio
SPL: Sound Pressure Level
SRT: Speech Recognition Threshold

## Acknowledgments

The authors thank Hansapanni Rodrigo for the statistical consultation.

