## Supplementary material for "Auditory deprivation during development alters efferent neural feedback and perception": Table 1

**Table 1. Summary of linear regression**

|  | | | | | | | | | | | |
| --- | --- | --- | --- | --- | --- | --- | --- | --- | --- | --- | --- |
| **Predictor** | **Estimate** | | | | **Standard Error** | | **t** | | **p** | | **Standardized Estimate** |
| Intercept ᵃ | |  | 11.84 |  | 1.48 |  | 7.98 |  | < 0.001 |  |  |
| Age | |  | –7.97 |  | 1.43 |  | –5.56 |  | < 0.001 |  | –0.48 |
| Absorbance | |  | –1.01 |  | 1.24 |  | –0.82 |  | 0.416 |  | –0.07 |
| SFOAE | |  | –9.39e−4 |  | 0.02 |  | –0.04 |  | 0.964 |  | –0.004 |
| Efferent inhibition | |  | –0.05 |  | 0.01 |  | –4.07 |  | < 0.001 |  | –0.36 |
| Group: | |  |  |  |  |  |  |  |  |  |  |
| Controls – OM group | |  | 0.96 |  | 0.37 |  | 2.58 |  | 0.012 |  | 0.48 |
| Sex: | |  |  |  |  |  |  |  |  |  |  |
| Female – Male | |  | –0.33 |  | 0.34 |  | –0.98 |  | 0.333 |  | –0.17 |
| ᵃ Represents reference level | | | | | | | | | | | |
